# Evaluation of Outdoor swimming courses as an intervention to refresh and revitalise NHS workers

**DOI:** 10.1101/2022.04.11.22273166

**Authors:** Pippa Pett, Heather Massey, Hannah Denton, Amy Burlingham, Mark Harper

## Abstract

**Background:** Frontline healthcare staff working in the National Health Service (NHS) have been, and continue to be, under a significant level of work related stress as a result of the COVID-19 pandemic. Long hours and greater clinical need have impacted negatively on work-life balance. The results of our preliminary studies indicate that outdoor swimming may be an effective treatment for anxiety and depression. We therefore hypothesised that the activity could improve symptoms of work-related burnout and stress in NHS workers. The primary objective of this study was to gather feedback from NHS staff participating in supervised swimming sessions that took place in an outdoor pool in London and the sea in Cornwall on the value and effectiveness of this initiative as they perceived it.

**Methods:** Following ethical approval (University of Portsmouth Science and Health research ethics committee SHFEC 2021-066), participants who had signed up to outdoor swimming courses provided by NHS Improvement in Cornwall and London were asked to give their consent to participate in an online survey. They were asked to complete them at three time-points: the week prior to, upon completion and six weeks after completion of the outdoor swimming course. As well as being asked for qualitative feedback, participants completed the Short Warwick-Edinburgh Mental Wellbeing Scale and the Copenhagen Burnout Inventory.

**Results:** 85 (63.9%) of the 133 Participants who signed up to outdoor swimming courses completed the first survey, 62 (49.6%) the second and 43 (35.5%) the third. 41 (33.8%) completed all three surveys. Overall, there was a 14.8% increase in wellbeing scores when comparing the scores before and after the courses which was statistically significant (p<0.0001, d= 1.02). Compared to scores before the course, the scores at its conclusion were reduced by 25%, 18% and 18% in personal, work-related and client-related burnout respectively. These burnout scores were significantly different for personal (P<0.0001) and work-related burnout (P=0.0018). Qualitative feedback was overwhelmingly positive with the effects being broadly divided into those relating to mood and physical health, the social aspects of the group activity, feelings of achievement and self-care and mindfulness.

**Conclusion:** This research suggests that the outdoor swimming activity, as a workplace intervention, can be an effective way of promoting staff wellbeing and reducing personal and work-related burnout. Further, formal trials of this intervention are justified.

## Introduction

CHILL CIC commissioned the University of Portsmouth to provide feedback from participants relating to outdoor swimming coaching sessions that were conducted for NHS staff as part of the NHS Improvement and wellbeing. The courses took place in London at Parliament Hill Lido and in Cornwall.

## Methods

### Participants and Procedures

Of the 133 Participants signed up to the course 121 agreed to be sent the survey link by e-mail. From the 121 e-mails that were sent 85 participants (70.2%) gave their consent to participate in the first survey, 62 (49.6%) in the second and 43 (35.5%) in the third. 41 (33.8%) completed all three surveys. Reminder e-mails were sent to all non-responders one week after the original survey was sent.

The outdoor swimming courses consisted of 8 sessions, each session was up to 1 hour in length. The sessions were supported by qualified and experienced open water swimming coaches and lifeguards.

Two courses were run in London – one in an open-air swimming pool which was relatively unchallenging for novice swimmers. The second was in the open water a reservoir.

### Surveys

Participants who had signed up to the outdoor swimming course were asked to give their consent to participate in each online survey. The study was ethically approved (University of Portsmouth Science and Health research ethics committee SHFEC 2021-066) and participants were free to leave the survey at any point.

The data collection consisted of three surveys which were conducted online using the JISC online survey platform. A reminder link was sent 7 days after the original survey request to participants who had not completed the survey. The survey time points were:

i. In the week prior to the outdoor swimming course,
ii. Upon completion of the outdoor swimming course and
iii. Six weeks after the completion of the outdoor swimming course.

The surveys included demographic information, asked for qualitative feedback about the course, ways in which it might be improved and validated measures of wellbeing and burnout outlined below:

1. Short Warwick-Edinburgh Mental Wellbeing Scale (SWEMWBS) (Haver et al 2015). The scale consists of seven items, responses ranged from 1 (none of the time) to 5 (all of the time) on a 5-point Likert scale. Individual indices were recorded; the total of the raw item-scores calculated and adjusted in accordance with Stewart-Brown et al. (2009) This shorter scale was preferred to the full version for brevity, but has a strong correlation with the original survey (*r* = .954). Cronbach’s alpha values for each of the subscales of the questionnaire ranged from 0.83 to 0.85 (Haver et al 2015).
2. Copenhagen Burnout Inventory (CBI, Kristensen et al 2005), Chronbach’s alpha 0.79-0.94 (Milfort et al 2008). The CBI consists of 19 items. It evaluates (i) personal related (6 items), (ii) work related (7 items) and (iii) client-related (6 items) burnout. Personal exhaustion refers to both physical and psychological fatigue that accumulates in a person during the day (*e*.*g*. ‘‘How often are you physically exhausted’’). Occupational exhaustion describes fatigue that is derived from work (*e*.*g*. ‘‘Do you feel worn out at the end of the working day’’). Client-related exhaustion depicts burnout as a consequence of interpersonal relationships with the clients (*e*.*g*. ‘‘Does it drain your energy to work with clients’’) (Kristensen *et al*., 2005). For each question participants scored:
  - 100% if noting an event or feeling occurred ‘always’,
  - 75% if the feeling was ‘often’,
  - 50% if the feeling occurred sometimes,
  - 25% if the feeling occurred ‘seldomly’ and
  - 0% if they ‘never’ experienced that feeling.

### Analysis

The data were anonymised prior to analysis.

#### Demographic data

The demographic data were descriptive data and have been recorded as frequencies.

#### Qualitative data analysis

This study, consistent with Braun and Clarke (2006), used thematic analysis in an open-ended way, to investigate how participants experienced the open water swimming courses. The authors (PP & HM) read and re-read transcripts in order to identify potential themes.

The second level of analysis involved reviewing these initial codes. At the third stage, quotes were selected to illustrate the overarching themes. Next, the authors reviewed themes prior to defining and naming them.

#### Quantitative analysis

The total of the raw item-scores for the SWEMWBS scale were calculated and adjusted in accordance with Stewart-Brown *et al*, (2009). The change in wellbeing level (low well-being 7–19 points, moderate wellbeing 20– 27 points and high well-being 28–35 points) (Stranges *et al*, 2014) and percentage with a positive or negative meaningful change in wellbeing (of 1–3 points) were reported in accordance with Shah *et al*. (2018). The total wellbeing scores were calculated for the whole group and then separately for the London and Cornwall group.

The total raw scores for the three parts of the burnout scales were calculated separately. Then mean scores were calculated for the whole group and London and Cornwall groups. All data were analysed descriptively, showing percentage changes in wellbeing and burnout, effect sizes calculated (Cohen’s d) to see if meaningful change had occurred and analysed statistically (using Wilcoxon sign rank tests) to see if there were differences that occurred other than chance. *d*=0.2 is interpreted as a small effect, 0.6 a moderate effect, 1.2 as a large effect and 2.0 or greater as a very large effect. A statistically significant difference is referenced as *P*<0.05.

## Results

### Course Attendance

The course attendance at the St Agnus courses in Cornwall was 69% and Parliament Hill Lido in London 63%. Tables 1-3 provide demographic data of the participants who attended the course.

**Table 1.** Gender, gender identity and age of participants.

| All data | Frequency | Mean (SD) |
| --- | --- | --- |
| **Gender** |  |  |
| Female | 73 |  |
| Male | 12 |  |
| Prefer not to say | 0 |  |
| Prefer to self-describe | 0 |  |
| **Gender identity same as sex assigned at birth?** |  |  |
| No | 1 |  |
| Yes | 82 |  |
| Prefer not to say | 1 |  |
| **Age (years)** |  | 40.5 (9.4) |

### Demographic data

From the original survey 30 participants were from London, 50 from Cornwall and 5 did not state their location.

### Qualitative analysis

Overall, the responses in the participant surveys after the course were positive in nature and whilst many individual themes emerged, they fell in to one of two broad categories:

- Effects specific to the activity of outdoor swimming.
- Effects of participating in an organised and structured group activity.

Some themes fell into both categories, particularly those pertaining to the positive mental health effects of participating.

#### Effects specific to the activity of outdoor swimming

##### Improvement in mood and physical health

Most of the participants felt a positive effect from the activity itself either in a mental or physical capacity. Specific themes which emerged were the improvement of stress management and improvement in mood both during and after the activity:

*‘Pressures at work continue to increase but I feel I am managing them better now I have the swimming to help de-stress and clear my head. Always feel more relaxed and happy once in the water and can leave work behind’*

A number of participants also cited improvements in sleep and physical fitness or weight loss from cold water swimming:

*‘Sleeping better and feeling less stressed. Lost weight’*

Some participants also felt beneficial effects from the feeling of their bodies adapting to the cold water both during and after the activity and were able to give specific examples of the physical changes they felt:

*‘Yes my breathing, mainly my breathing rate overall has reduced, meaning I’m in parasympathetic system. reducing my rest heart and helping me to lose weight, cause my cortisol level to reduce’*

Many participants also felt a positive effect and an improvement in confidence from regularly swimming and being outdoors. These responses were found in greater number in the Cornwall group who felt that the act of going in the sea itself had a positive effect. The same themes were found in the London (Lido) group but in fewer numbers:

*‘The sea is invigorating and keeps you in the HERE and NOW. it is a very grounding thing to do to just freeze your butt off and BE in the moment outdoors away from all the faff of the NHS and be in nature’ (Cornwall)*

*‘I feel more confident in my swimming and have got back into a routine of swimming. I also feel like it helps my breathing and my mood*.*’ (London)*

#### Effects of participating in an organised and structured group activity

##### Positive effect of a group activity

Many of the participants especially enjoyed participating in a social group activity and some also enjoyed the aspect that most people were new to the activity and they shared a common background with them (NHS):

*‘The social aspect and meeting others who are as nervous as me!’ (London)*

*‘I enjoyed getting together with like-minded individuals. the experience of cold water swimming has been fabulous*.*’*

*‘Meeting new people who have also been working in the NHS throughout the pandemic’ (Cornwall)*

Some participants felt that having a group activity to go to encouraged them to continue with it more than if they had been exercising in isolation:

*‘Great to meet new people and encourage outdoor swim every week. Definitely would have not gone every week if it wasn’t for the group’*

##### Being coached and trained by experts to learn a new skill

Responses that mentioned the presence of trainers/coaches were all positive and many of the responses valued the opportunity to learn a new skill with the support of a professional:

*‘I enjoyed getting involved in a new experience, that was organised and all we had to do was to learn new skills, new information’ (London)*

*‘Excitement of being in huge waves with support of the teacher. ‘(Cornwall)*

Some felt that the activity was made more accessible to them due to the structure of the programme and a levelling effect of having participants of all abilities:

*‘Yes, I particularly liked being on the beach/in the sea in a small group of people and it not mattering that I didn’t have any experience of what I was doing (I’m in my 40s so if I take up new activities I always feel awkward because other people my age know what they’re doing)’*

##### Sense of achievement of participating in a new and challenging activity

For many participants they felt that participating in an activity of this challenging nature gave them a sense of achievement and as a result this improved their confidence and overall enjoyment of the activity:

*‘Yes I feel more confident going into the water alone and in general. when I was doing it weekly i found myself looking forward to it! It was relaxing and an achievement*.*’*

Others felt that participating in and completing the activity gave them a sense of achievement:

*‘A feeling of achievement and completing something*.*’*

##### Self-care and mindfulness

Many of the participants felt that participating in an activity which was organised and structured forced them to take time for themselves. There was a feeling of ‘protected time’ in some of the responses in having a commitment to attend the activity:

*‘Good at keeping boundaries at work as I had something to get to after work. Felt easier to put these boundaries in place and not stay late*.*’*

Others specifically cited the activity as an act of ‘self-care’ or ‘me time’:

*‘I can’t change my work and the pressure there or in my home life but I can change what I do for myself and how I do it. It is one of my few acts of self-care’ (London)*

‘I feel it is time for me and that is special.’ (Cornwall)

##### Overall satisfaction with the activity

In general, the majority of participants felt that the activity had a positive effect and the response to the question ‘Can you describe anything about the activity that you have not liked?’ was mostly ‘no’ or ‘nothing’. Some participants mentioned things they would change about structure or location for example distance to locations, timings and parking was challenging for some of the participants:

*‘Beach choice and session times. It was very difficult to park throughout the summer sessions*.*’*

Most of the participants expressed a desire for the course to continue to run and for others to have access to the course. Some specifically mentioned it in relation to the NHS the people who worked for the organisation:

‘None at all - more sessions so others can benefit! My colleagues have all worked so, so hard over the past couple of years and I ‘hope they will be able to access this.’

*‘introduce across more trusts. it is the best thing our trust has done for us*.*’*

One participant attended both of the London courses and felt that moving from an open-air lido to a reservoir after only 8 sessions was a large step in skill and competence level. They felt this highlighted important issues in access to sport and exercise in general and this activity in particular.

### Quantitative survey data

#### SWEMWBS – Wellbeing

##### All data

Table 4 provides all the SWEMWBS data across all participants in London and Cornwall. For separated data please see appendix 1. Table 2 the breakdown of the values by wellbeing level, age and gender.

**Table 2.** The ethnic group of the participants

|  |  |
| --- | --- |
| **Ethnic group** |  |
| *Asian/Asian British* |  |
| Indian | 2 |
| Bangladeshi | 0 |
| Chinese | 0 |
| Pakistani | 0 |
| Other Asian background | 0 |
| *Black/ African/Caribbean/Black British* |  |
| African | 2 |
| Caribbean | 1 |
| Any other Black/African/Caribbean background | 1 |
| *Mixed/Multiple ethnic groups* |  |
| Black Caribbean and White | 9 |
| Black African and White | 0 |
| Asian and White | 0 |
| Black Caribbean and Black African | 4 |
| *White* |  |
| English/Welsh/Scottish/Northern Irish/British | 49 |
| Irish | 0 |
| Traveller | 0 |
| Any other White background,<br>please describe | 36 |
| Arab | 0 |
| Other ethnic group | 0 |

**Table 3.**
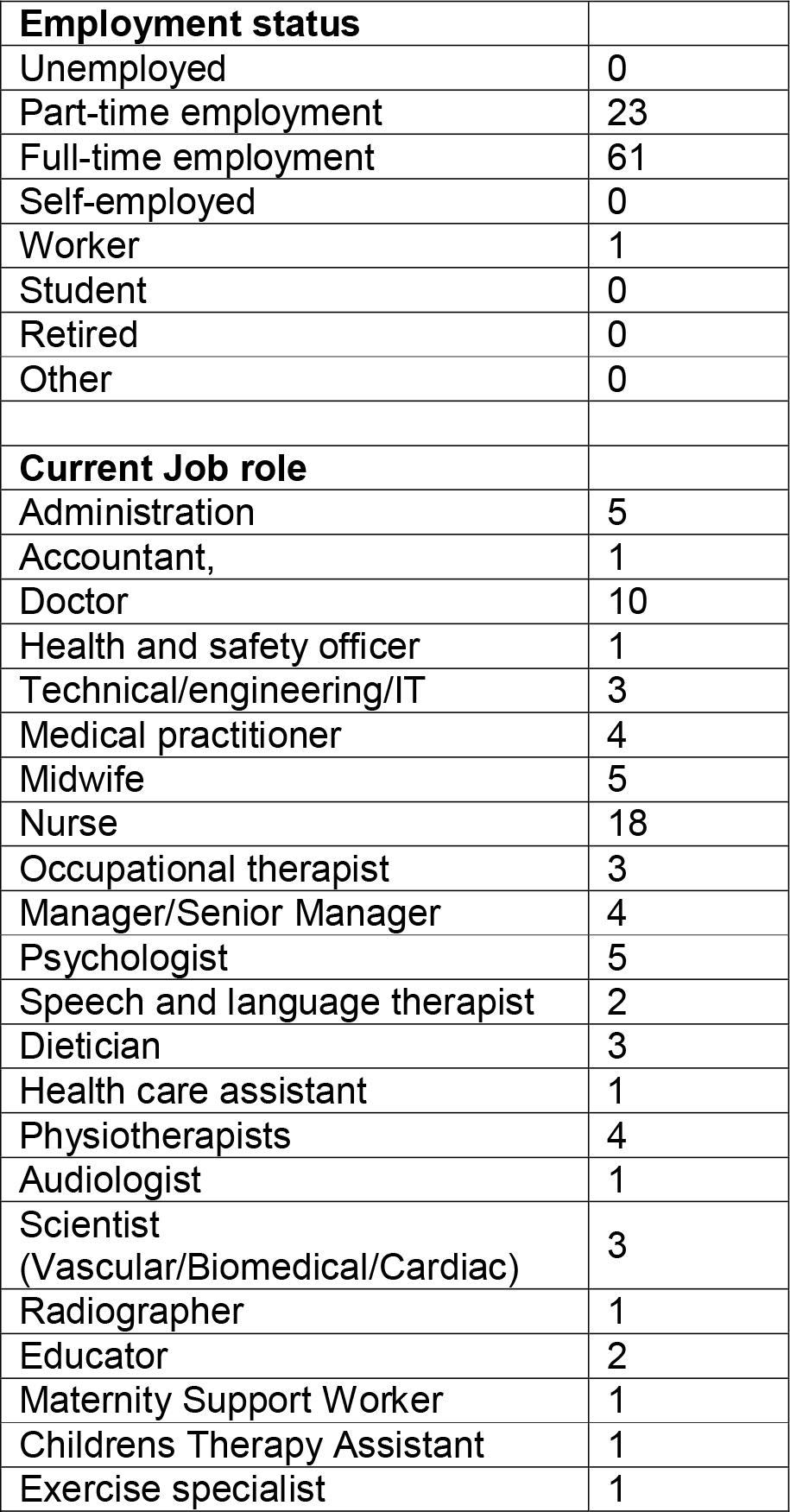
Employment status of participants

**Table 4.** Mean and standard deviation of all SWEMWBS wellbeing scores before the course, immediately after the course and a 6 week follow up

| All data | I've been feeling optimistic about the future | I've been feeling useful | I've been feeling relaxed | I've been dealing with problems well | I've been thinking clearly | I've been feeling close to other people | I've been able to make up my own mind about things | Total |
| --- | --- | --- | --- | --- | --- | --- | --- | --- |
| **Before course (n=85)** |  |  |  |  |  |  |  |  |
|  | 3.3 (0.8) | 3.4 (0.7) | 2.7 (0.8) | 3.2 (0.8) | 3.2 (0.8) | 3.2 (0.9) | 3.4 (0.9) | 22.4 (3.9) |
| **After course (n=62)** |  |  |  |  |  |  |  |  |
|  | 3.8 (0.8) | 3.9 (0.7) | 3.4 (0.6) | 3.6 (0.7) | 3.6 (0.6) | 3.8 (0.8) | 4.1 (0.7) | 26.3 (3.6) |
| **Follow up (n=43)** |  |  |  |  |  |  |  |  |
|  | 3.8 (0.7) | 3.9 (0.6) | 3.5 (0.8) | 3.6 (0.7) | 3.6 (0.7) | 3.7 (0.8) | 4.0 (0.7) | 26.2 (3.7) |

**Table 5.** Breakdown of SWEMWBS scores by wellbeing level, age and gender and the percentage having a meaningful change in wellbeing in participants who completed all three surveys. *Statistically significantly different from before the course *P<0*.*0001*

|  |  | Survey time point |  |  | Comparisons |  |  |
| --- | --- | --- | --- | --- | --- | --- | --- |
|  |  | Before course | After course | Follow up | Before v after | before v follow up | after v follow up |
| Total no. of responses |  | 85 | 62 | 43 |  |  |  |
| % Low wellbeing |  | 53% | 12% | 14% |  |  |  |
| % Moderate wellbeing |  | 42% | 76% | 77% |  |  |  |
| % High wellbeing |  | 5% | 12% | 9% |  |  |  |
| Mean score |  | 22.4 | **26.3\*** | **26.2\*** | 3.9 | 3.8 | 0.1 |
| Standard deviation |  | 3.9 | 3.6 | 3.7 | 4.4 | 4.6 | 2.8 |
| Effect size d= |  |  |  |  | 1.02 | 1.03 | 0.02 |
|  |  |  |  |  | moderate | moderate | none |
| Mean Score |  |  |  |  |  |  |  |
| By age | 25-39 | 22.6 | 27.1 | 28.0 | 4.5 | 5.4 | 0.9 |
|  | 40-54 | 22.0 | 25.5 | 25.3 | 3.5 | 3.3 | -0.3 |
|  | 55-64 | 23.5 | 26.0 | 25.8 | 2.5 | 2.3 | -0.2 |
| By gender |  |  |  |  |  |  |  |
|  | Male | 24.1 | 28.5 | 25.5 | 4.4 | 1.4 | -3.0 |
|  | Female | 22.2 | 25.9 | 26.3 | 3.7 | 4.1 | 0.4 |
| N with a meaningful positive change (%) |  |  |  |  | 30 | 22 | 6 |
| N with a meaningful negative change (%) |  |  |  |  | 2 | 2 | 4 |
| a meaningful positive change (%) |  |  |  |  | 54.5 | 53.7 | 18.2 |
| a meaningful negative change (%) |  |  |  |  | 3.6 | 4.9 | 12.1 |

Overall there was a 14.8% increase in wellbeing scores when comparing the scores before compared to after, this increase in scores was statistically significantly different and had a moderate effect (p<0.0001, d= 1.02, Table 2).

In London, total wellbeing scores were increased by 9.1% following the course and remained elevated in follow up.

In Cornwall, total wellbeing scores were increased by 18.2% following the course and again remained elevated in follow up.

##### Results from those who completed all surveys

Six weeks after the conclusion of the course, wellbeing remained elevated compared to beforehand (p<0.0001, d=1.03). Wellbeing was similar at follow-up when compared to the end of the course (p=0.673 d=0.02) indicating that the intervention had a persistently positive effect.

A high percentage of participants initially had a low wellbeing score before the course yet moved into the moderate wellbeing category following it and remained there in follow up (Table 2).

All age groups showed a rise in wellbeing scores after the course which appeared to be maintained at follow-up. However, when broken down by gender, while females maintained their wellbeing levels, males showed a large increase in SWEMWBS scores at the end of the course but this decreased appreciably at follow-up (Table 2)

#### CBI - Burnout

##### All data

Table 6 provides all the burnout data from all participants. For data separated by location please see appendix 1.

**Table 6.**
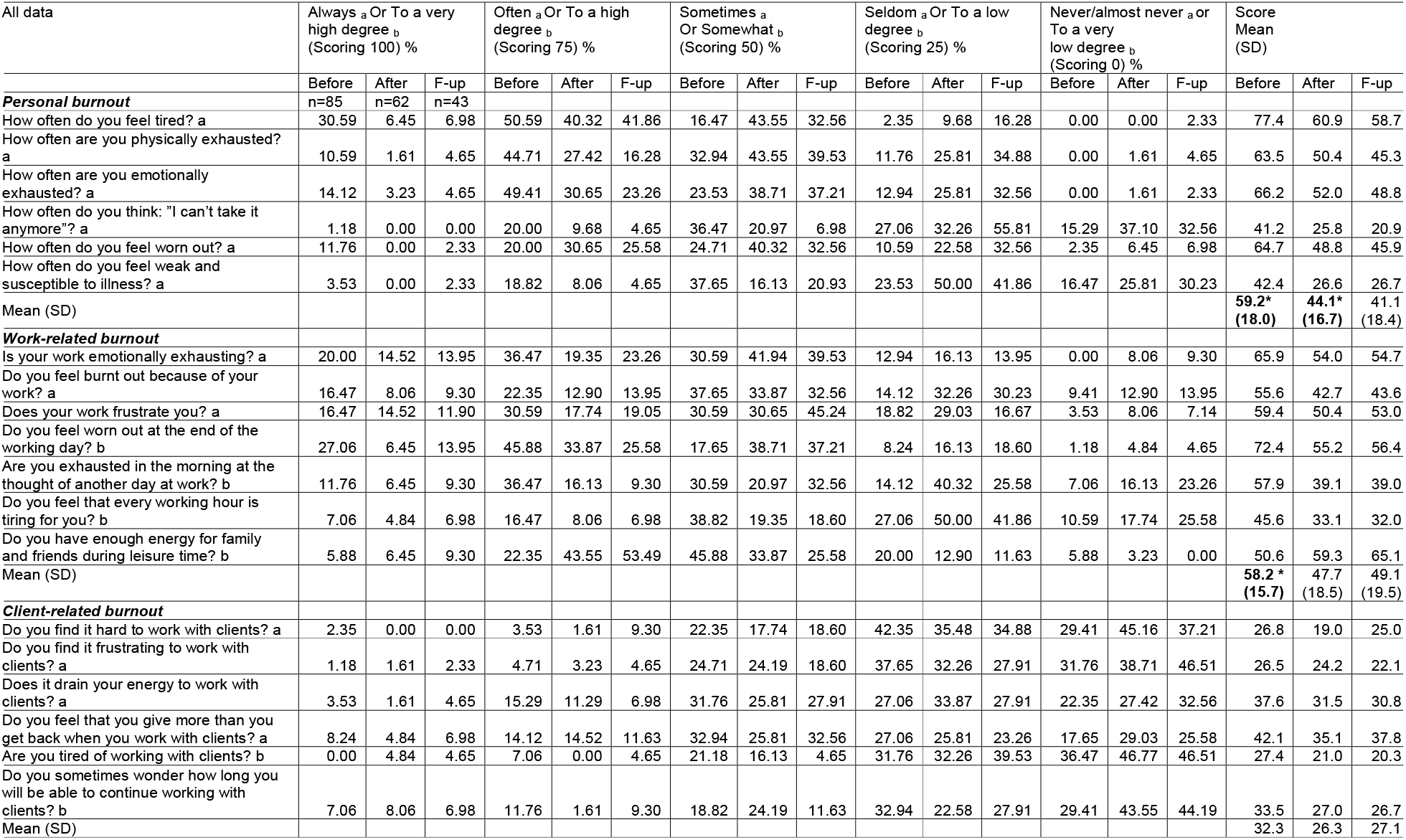
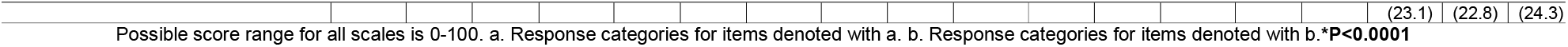
Percentage of values expressed for all the CBI data collected in all participants in London and Cornwall. For data separated by location please see appendix 1.

##### Overall scores,

There are three categories of burnout, personal, work- and client-related burnout. Compared to scores before the course, the scores at its conclusion were reduced by 25%, 18% and 18% in personal, work-related and client-related burnout respectively. These burnout scores were significantly different for personal (P<0.0001) and work-related burnout (P=0.0018, Table 6).

Personal burnout was also reduced between the start of the course and the six-week follow up (p<0.0001. Neither personal burnout or work-related burnout differed significantly between the post-course and follow up surveys (Personal p=0.2988 and work-related p=0.8751 burnout, Table 6)

Client-related burnout did not differ at any time point; when comparing the surveys before and after the course (p=0.3152), before and in follow up (p=0.2337) and after and in follow up (p=0.9679). Client burnout scores were low at all survey points.

In London, burnout scores reduced between the start and end of the course by 23.8% (personal burnout), 13.4% (work-related burnout) and 19.3% (client-related burnout). And in Cornwall burnout scores were also reduced between the surveys conducted before and after the course, by 28.4% (personal burnout), 24.7% (work-related burnout) and 26.0% (client-related burnout).

## Discussion

The primary objective of this study was to gather the views of NHS staff participating in the outdoor swimming sessions taking place at Parliament Hill Lido in London and in Cornwall on the value and effectiveness of this initiative. This document provides their feedback, and provides opportunities to improve the offering to NHS and, possibly, other groups of employees in the future.

The narrative feedback from participants in this course is overwhelmingly positive and the majority of those involved spoke only of benefits both physically and mentally.

It is clear that the activity itself had a beneficial effect for most of those participating but some other themes emerged which also showed that an organised activity, supported by professionals had, in itself, beneficial effects. Many people in the study cited the social aspect as being one of the most valued parts of the course.

Often during extremely challenging and busy times in a workplace, despite the shared experience, there can be a feeling of disconnection with colleagues. There is a need to ‘keep going’ and normalise an extreme situation which denies colleagues the space and opportunity to process it. The fact that so many people found the social side of the activity to be beneficial, suggests that offering it to more people within the NHS could help to build connections with colleagues and create the feeling of community which has been lost during a pandemic where the working environment has been radically altered by social distancing, PPE and the sheer volume of work preventing people sitting down together iin an organisation that was already under immense pressure.

Other important themes which emerged from the qualitative data were around inequalities in and access to sports and exercise. For some it was a feeling that if you aren’t already proficient in an activity then there is no easy way to access it, especially with advancing age. The nature of the course itself with the involvement of people of all ages and abilities seemed to remind some participants that regardless of the stage of life you are at, you can still try new things and develop new skills. The idea of developing a new skill was one of the positive effects that participants spoke of and the way that the course was structured seemed to give participants the feeling of security and safety that they needed to do this.

The challenge of engaging an ethnically and socio-economically diverse population was highlighted by one participant who noted a difference between the pool and open-water courses in London, emphasising the ongoing need to be vigilant with respect to maintaining accessibility. This also highlights the value of the less ‘wild’ environment of the outdoor pool and could be further mitigated by the introduction of “CHILL +” courses to provide a follow-on from the first lido course to bridge this gap in water competence and the participants’ feeling of security.

There is no question that the pressures on the NHS have been increasing over the past few years which have made things exceptionally challenging for everyone who works within it. The pandemic has increased these pressures further and have been exacerbated by the additional non-work-related stresses generated by COVID-19-related societal challenges. In general, working in Healthcare is thought of as a vocation more than as a way to pay the rent. With that comes a certain dedication which, whilst beneficial to the system in the short term, can be detrimental to the individual over time. It is, therefore, essential to protect and provide for the mental and physical health of staff, as this will reap benefits to the whole of society in the long term.

For many people working in healthcare roles, the thought of ‘me time’ or ‘self-care’ is a complex concept. Healthcare professionals are trained to ‘put the patient first’ and so the thought of self-care activity can inspire guilt and can feel almost juxtaposed to professional values. Instead healthcare professionals may be more prone to indulging in negative coping mechanisms which can cause harm in the long term. For this group of people it seems much more intuitive to take that first step towards developing healthy coping mechanisms in an activity which is organised for them by their Trusts, giving the sense of it being ‘rubber stamped’ with the additional draw of feeling that you are also giving your time as part of a study.

It is understandable that running the sessions in this context would draw the people who needed it most (those who are arguably least likely to participate in self-care) to be involved and to open the door to them understanding the benefits of this protected time with a view to developing healthier coping mechanisms in the future. As one participant put it *‘it is the best thing our Trust has ever done for us*’. This suggests that as well as the physical, mental and social benefits to the activity there could also be an opportunity for Trusts across the country to show care for their employees in a relatively simple way with the potential to have huge positive effects on their workforce.

The days when food, accommodation and other benefits were provided to healthcare workers are gone and although there are understandable reasons why these services were cut, there is a feeling that the ‘care for those who care for you’ feeling has also gone as well.

In theory healthcare work attracts a certain type of person. For the most part those people are caring, hardworking and dedicated and we need to create a structure which ensures that staff retain those qualities rather than losing them or forcing them to question why they chose the profession in the first place. We have a duty to protect and support the people who dedicate their professional lives to the care of our friends and families, to ensure that they are both physically and mentally strong in order to continue to do this. To echo the sentiments of one participant, we can’t change the pressure of work or home life but what we could do is offer small gestures to give people greater ability to manage them.

The quantitative data reflects many of the views expressed. Of the final survey respondents, 99% had continued outdoor swimming in between the course and follow up survey, 50% of those had been swimming at least once a week. The participants also indicate that their wellbeing increased and burnout scores were reduced following the outdoor swimming course and in most cases were maintained in the six-week follow-up period.

The improved wellbeing scores showed that initially over 50% of the staff surveyed had low wellbeing, following the course wellbeing had improved with more than 75% having moderate wellbeing and a small increase in those with high wellbeing. Similar wellbeing improvements have been seen in previously published research into open water swimming interventions with the general public (Massey et al 2020).

Both personal and work-related burnout before the start of the courses were high and higher than previous studies of healthcare workers and uniformed services (Kristensen *et al* 2005, Milfont *et al* 2008, Santa Maria *et al* 2019). Both personal and work-related burnout were reduced after the course and in follow up, but remained elevated. On the other hand, ‘client-related’ burnout was low to moderate as several participants commented that the burnout they experienced was not centred around patients. Participants felt proud to be able to help patients and did not therefore see them as a source of burnout.

There may be other contributing factors to the reduction in burnout and improved wellbeing, these may be unrelated to the outdoor swimming course, such as reduced immediate pressure from COVID infection rates and hospitalisations, increased opportunities for holidays and fewer lockdown restrictions and the change in the season towards summer. These cannot be discounted as contributing to the results.

Outdoor swimming as a workplace intervention may not be for everyone. Therefore, a range of activities for staff that provide opportunities to learn new skills, be outdoors, be physically active and an opportunity to meet colleagues in a social environment should be considered. In addition, continued efforts are needed to ensure that inclusion is carefully considered at every stage.

## Conclusion

This research suggests that the outdoor swimming activity, as a workplace intervention, can be an effective way of promoting staff wellbeing and reducing personal and work-related burnout. The comments received indicate that the outdoor swimming courses offered a positive experience both in itself and through learning a new skill and by providing a highly-valued opportunity to socialise and meet a new group of people who work in the NHS and have been through similar experiences. Further, formal trials of this intervention are justified.

## Data Availability

All data produced in the present study are available upon reasonable request to the authors

## Acknowledgements

This survey could not have been completed without the support of the participants and the following people, Swim coaches Keri-Anne Payne, Omie Dale, Mike Morris CHILL CIC, Laura Emerson NHS Improvement London, and Anna Dalziel NHS Improvement Cornwall.

### Appendix 1. Qualitative and Quantitative data

#### New experience/skills

the experience and training and social aspect.’

‘Meeting new people from the NHS. Learning new skills re swimming’

‘Creating a really safe group of NHS colleagues that felt safe to push myself in’

‘I enjoyed getting involved in a new experience, that was organised and all we had to do was turn new skills, new information in regards to’

‘Trying something new, enjoying it, meeting new people, time for me, the feel of being in the sea and everything else just has to wait, even for just 20mins’

‘Getting outdoors, meeting new people and doing a new exciting activity’

‘Yes, I particularly liked being on the beach/in the sea in a small group of people and it not mattering that I didn’t have any experience of what I was doing (I’m in my 40s so if I take up new activities I always feel awkward because other people my age know what they’re doing)’

‘Learning to swim in the sea’

‘I loved how it made me feel, meeting other people and learning more about sea safety.’

‘Being with others. Meeting new people, who I don’t know but work in the same hospital in different departments. Social Aspect. Getting to learn different things about the ocean, the tides, etc from the lifeguards. And overcoming my fears of swimming in the open ocean.’

‘Learning to sea swim

‘I can now enjoy swimming under water which was my goal :)’

‘I have more knowledge about the sea.’

‘opened my mind to something new’

#### Being outdoors/in sea/active

Being outside, the community that developed, lovely supportive atmosphere’

‘Getting outdoors in the evening after work’

‘The swimming, having space in the lido to ourselves, the other people there’

‘The opportunity for a free swim outdoors after work and at the beginning the chance of being coached. Nice to catch up and meet others too’

‘Opportunity to regularly swim outside. Good quality of the teaching’

‘wonderful to meet other NHS workers and great to get outside’

‘Trying something new, enjoying it, meeting new people, time for me, the feel of being in the sea and everything else just has to wait, even for just 20mins’

‘Increased confidence and enjoyment of sea swimming’

‘Getting outdoors, meeting new people and doing a new exciting activity’

‘Yes, I particularly liked being on the beach/in the sea in a small group of people and it not mattering that I didn’t have any experience of what I was doing (I’m in my 40s so if I take up new activities I always feel awkward because other people my age know what they’re doing)’

‘Excitement of being in huge waves with support of the teacher.’

‘The feeling of freedom, the women I met and the feeling of accomplishment.’

‘Learning to swim in the sea’

‘Sense of achievement from being nervous at the beginning to excitement about getting in the water.’

‘Great to meet new people and encourage outdoor swim every week. Definitely would have not gone every week if it wasn’t for the group’

‘The fact that it seems to have recalibrated my system. Hard to put into words but I feel like I have found a bit of my old self, I love the sea again, I’m not in battle with it, I’m swimming at least twice a week at the moment and Im much much happier.’

‘Helped me feel more in the moment and in touch with nature. I always feel better after being in the sea both mentally and physically.’

‘With it being a group activity, it made me go, even if i wasn’t always ‘feeling it’. I loved the feel of the cold water as it engulfed me.’

‘Social aspect, feeling of the water on skin, being outside and the views’

‘the social aspect as well as being outdoors. Loved the feeling of getting used to the temperature of the water. the progression each week’

‘will be more confident swimming outdoors in cold water and will go in sea more’

‘I am much more confident in the water and in my swimming ability’

‘Yes I am doing more exercise’

‘I can now enjoy swimming under water which was my goal :)’

‘I feel more confident in my swimming and have got back into a routine of swimming. I also feel like it helps my breathing and my mood.’

‘My swimming technique has improved. I’m regularly swimming outdoors.’

‘I am doing more swimming outdoors.’

‘Much more confident as a swimmer and will now be exploring more outdoor swimming’

‘I am more confident to swim in the sea and am not so worried about jelly fish now that Sophie has reassured us that the sting of most types are no worse than a stinging nettle and some have no sting at all!’

‘Really want to swim each day’

‘I now routinely sea swim/paddle more regularly’

‘I go outdoor swimming at least once a week with some lovely ladies who did the course too.’

‘Yes, I now swim in the sea several times a week. It gives me peace, calm and a sense of ok-ness with the world’

‘Yes - doing more outdoor swimming’

‘I now regularly participate in cold water swimming.’

‘I’m in general more active and more motivated to continue to be more active - doing more yoga, interval training and carb cycling/healthy eating with husband. I’m less stressed.’

‘Yes, I am now a regular sea swimmer’

‘More motivation to get in the sea. I encourage others to try it too now.’

‘Yes I feel more happy overall, I think this is that I’m now doing more outdoors based activity each week’

‘Embrace the cold, feeel the fear and do it anyway’

#### Being coached/trained

‘the experience and training and social aspect’

‘The instructors were very kind and helpful’

‘The opportunity for a free swim outdoors after work and at the beginning the chance of being coached. Nice to catch up and meet others too’

‘Opportunity to regularly swim outside. Good quality of the teaching’

‘Excitement of being in huge waves with support of the teacher.’

‘loved every minute of it. The instructor was great and the group lovely’

‘The confidence boost that it gave me to just do it! The Trainers were amazing. The distance’

‘The Chill UK team were excellent’

#### Social interaction/community/solidarity

‘the experience and training and social aspect’

‘Meeting new people from the NHS. Learning new skills re swimming’

‘Being outside, the community that developed, lovely supportive atmosphere’

‘Creating a really safe group of NHS colleagues that felt safe to push myself in’

‘The swimming, having space in the lido to ourselves, the other people there’

‘The peer group element and inclusive nature of the sessions’

‘Meeting a range of people who are from all over the NHS in London’

‘The opportunity for a free swim outdoors after work and at the beginning the chance of being coached. Nice to catch up and meet others too’

‘wonderful to meet other NHS workers and great to get outside’

‘The social aspect and meeting others who are as nervous as me!’

‘Trying something new, enjoying it, meeting new people, time for me, the feel of being in the sea and everything else just has to wait, even for just 20mins’

‘Getting outdoors, meeting new people and doing a new exciting activity’

‘Meeting new friends Increasing confidence in the water’

‘The feeling of freedom, the women I met and the feeling of accomplishment.’

‘Meeting new people who have also been working in the NHS throughout the pandemic’

‘I enjoyed getting together with like-minded individuals. the experience of cold water swimming has been fabulous.’

‘With it being a group activity, it made me go, even if i wasn’t always ‘feeling it’. I loved the feel of the cold water as it engulfed me.’

‘loved every minute of it The instructor was great and the group lovely’

‘Time to myself Meeting new people Challenging myself’

‘I loved how it made me feel, meeting other people and learning more about sea safety.’

‘Social aspect, feeling of the water on skin, being outside and the views’

‘Being with others. Meeting new people, who I don’t know but work in the same hospital in different departments. Social Aspect. Getting to learn different things about the ocean, the tides, etc from the lifeguards. And overcoming my fears of swimming in the open ocean.’

‘Great to meet new people and encourage outdoor swim every week. Definitely would have not gone every week if it wasn’t for the group’

‘making more time to swim with peers and group’

‘Feel more connected to others encouraged to attend the pool, even though I have a season ticket I think I’d have stopped without the group’

‘Making time for me to go swimming at least once a week, still meeting new people, calmer and more confident’

‘I go outdoor swimming at least once a week with some lovely ladies who did the course too.’

‘I am now meeting with others and swimming on a regular basis. We hope to continue throughout the winter months too.’

‘I became covid positive at the end of the sessions and sadly haven’t been able to continue due to symptoms. I am really keen to start meeting other people from the other courses’

#### Confidence

Increased confidence and enjoyment of sea swimming’

‘Improved my confidence in the sea hugely and after the course still swimming regularly’

‘Meeting new friends. Increasing confidence in the water’

‘It has built my confidence - I am much less fearful about everything as a result’

‘confidence boost’

‘The confidence boost that it gave me to just do it! The Trainers were amazing The distance’

‘will be more confident swimming outdoors in cold water and will go in sea more’

‘I am much more confident in the water and in my swimming ability’

‘I feel more confident in my swimming and have got back into a routine of swimming. I also feel like it helps my breathing and my mood.’

‘I feel more confident in managing cold water immersion and know how my body responds’

‘Much more confident as a swimmer and will now be exploring more outdoor swimming’

‘Making time for me to go swimming at least once a week, still meeting new people, calmer and more confident’

‘I am more confident to swim in the sea and am not so worried about jelly fish now that Sophie has reassured us that the sting of most types are no worse than a stinging nettle and some have no sting at all!’

‘Yes I feel confident to make changes and be more assertive in my every day life.’

‘I have more confidence in myself.’

‘confident in doing sea swimming and other outdoor water activities’

‘More confidence in the water.’

‘yes I am more confident ion open water’

‘Yes, more confidence to go swimming on my own and I do it more regularly as I realise the benefits’

‘I’m braver with cold water/sea - rather than quit & get out I now stay in knowing I’ll adapt to the temperature’

‘Yes I feel more confident going into the water alone and in general. when I was doing it weekly i found myself looking forward to it! It was relaxing and an achievement.’

‘Gave me the confidence that i can get into cold sea water and it feels great.’

‘I have the confidence to go in the water by myself and the curiosity to learn more about the moon & tides.

As a menopausal woman, I feel this has given me some confidence back.’

#### Sense of achievement/accomplishment

‘The feeling of freedom, the women I met and the feeling of accomplishment.’

‘Sense of achievement from being nervous at the beginning to excitement about getting in the water.’

‘Time to myself Meeting new people Challenging myself’

‘Happier in myself for sticking with it’

‘I can now enjoy swimming under water which was my goal :)’

‘Yes I feel more confident going into the water alone and in general. when I was doing it weekly i found myself looking forward to it! It was relaxing and an achievement.’

This has been brilliant and made a huge difference to me its been a positive experience’

‘I’m glad I signed up to this, as I was nervous about doing this (I hadn’t swum since getting COVID and long-COVID). Thank you to the team for arranging this’

‘I am really grateful for this opportunity, it has been amazing’

‘Thank you for the opportunity’

‘It was an amazing experience.’

#### Self-care and mindfulness

‘Trying something new, enjoying it, meeting new people, time for me, the feel of being in the sea and everything else just has to wait, even for just 20mins’

‘The lovely feeling of achievement’

‘Time to myself Meeting new people Challenging myself’

‘I loved having a set time for me. This meant I had to leave work on time’

‘making more time to swim with peers and group’

‘Making time for me to go swimming at least once a week, still meeting new people, calmer and more confident’

‘being conscious that i can make the time to get in the sea and that it is worth it when I make the effort’

#### Positive physical effects

‘The fact that it seems to have recalibrated my system. Hard to put into words but I feel like I have found a bit of my old self, I love the sea again, Im not in battle with it, Im swimming at least twice a week at the moment and Im much much happier.’

‘Helped me feel more in the moment and in touch with nature. I always feel better after being in the sea both mentally and physically.’

‘Social aspect, feeling of the water on skin, being outside and the views’

‘yes my breathing, mainly my breathing rate overall has reduce, meaning I’m in parasympathetic system. reducing my rest heart and helping me to lose weight, cause my cortisol level to reduce’

‘I feel more confident in my swimming and have got back into a routine of swimming. I also feel like it helps my breathing and my mood.’

‘I feel more confident in managing cold water immersion and know how my body responds’

‘My swimming technique has improved. I’m regularly swimming outdoors.’

‘Sleeping better and feeling less stressed. Lost weight’

‘think so, I’m planning to continue sea-swimming but have ended up taking a few weeks out due to other commitments (holiday away, visting family etc). My mood was definitely better during the sessions and I did sleep better.’

‘i feel that this type of activity can be very beneficial to other population and will be really interesting to see the positive adaptation the body experiences. how can we use these changes to help with prevention, rehabilitation and pre-habilitation etc’

‘No - only to say that the feeling of immersion in cold water is something I would never have believed I would grow to love. It clears away anxiety, stress and any feelings of overwhelm. I am a much happier person because of sea swimming.’

#### Improved stress management

‘yes my breathing, mainly my breathing rate overall has reduce, meaning I’m in parasympathetic system. reducing my rest heart and helping me to lose weight, cause my cortisol level to reduce’

‘Sleeping better and feeling less stressed. Lost weight’

‘Yes, I now swim in the sea several times a week. It gives me peace, calm and a sense of ok-ness with the world’

‘I’m in general more active and more motivated to continue to be more active - doing more yoga, interval training and carb cycling/healthy eating with husband. I’m less stressed.’

‘No - only to say that the feeling of immersion in cold water is something I would never have believed I would grow to love. It clears away anxiety, stress and any feelings of overwhelm. I am a much happier person because of sea swimming.’

#### Challenges or additional stressors

‘No as I then got into managing school holidays and slipped out of the habit so need to get back in to it now’

I would love to be able to take part in the whole course t some point if location was nearer to me.’

#### Happier/more positive outlook/more relaxed

‘The fact that it seems to have recalibrated my system. Hard to put into words but I feel like I have found a bit of my old self, I love the sea again, Im not in battle with it, Im swimming at least twice a week at the moment and Im much much happier.’

‘I feel more confident in my swimming and have got back into a routine of swimming. I also feel like it helps my breathing and my mood.’

‘I think so, I’m planning to continue sea-swimming but have ended up taking a few weeks out due to other commitments (holiday away, visting family etc). My mood was definitely better during the sessions and I did sleep better.’

‘Yes I feel confident to make changes and be more assertive in my every day life.’

‘Yes, I now swim in the sea several times a week. It gives me peace, calm and a sense of ok-ness with the world’

‘my mental health has improved immensely. i am more motivated, less scared.’

‘Yes I feel more happy overall, I think this is that I’m now doing more outdoors based activity each week’

‘Yes I feel more confident going into the water alone and in general. when I was doing it weekly i found myself looking forward to it! It was relaxing and an achievement.’

‘No - only to say that the feeling of immersion in cold water is something I would never have believed I would grow to love. It clears away anxiety, stress and any feelings of overwhelm. I am a much happier person because of sea swimming.’

#### Inequalities in access to sport

‘Yes - the process for recruitment on to the 2nd course (open water swim at West reservoir) threw away all of that support and confidence by not having Parliament Hill as a feeder course. It decide to “pull the name out of a hat” because so many people had applied. This approach fundamentally re-enforces structural discrimination. Of course, this 2nd course is full of the usual suspects - skinny white woman who are already doing a lot of swimming as exercise. At the parliament hill course half of us turned up without googles coz we weren’t swimmers and didn’t think about kit. At the West reservoir, they are all showing up with the outdoor swimming ponchos on session #1. The original is course wasn’t about swim capability - it was about MH and fitness. Something the second course hasnt recognised’

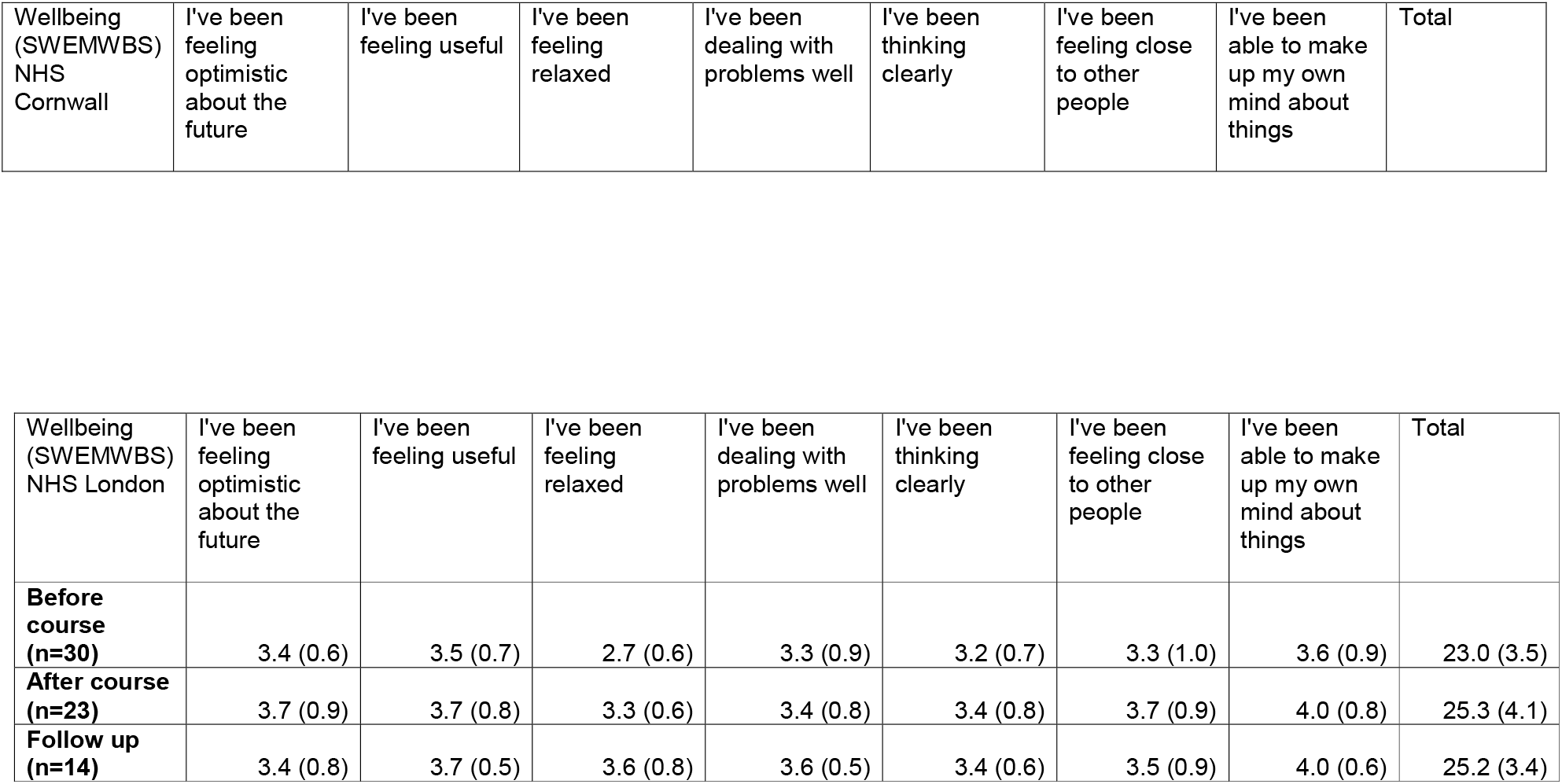
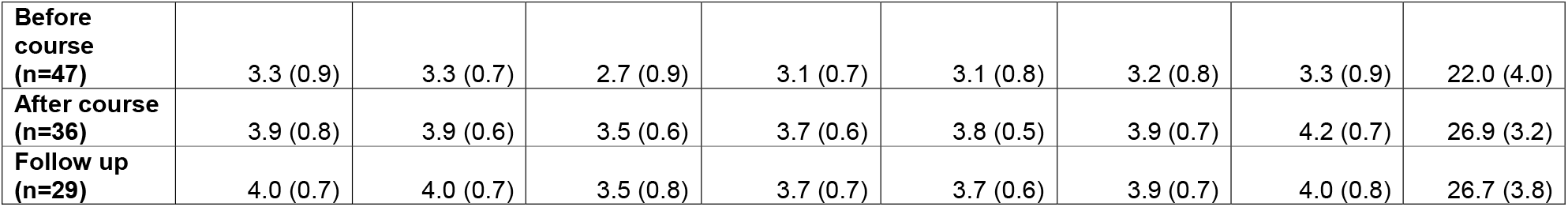
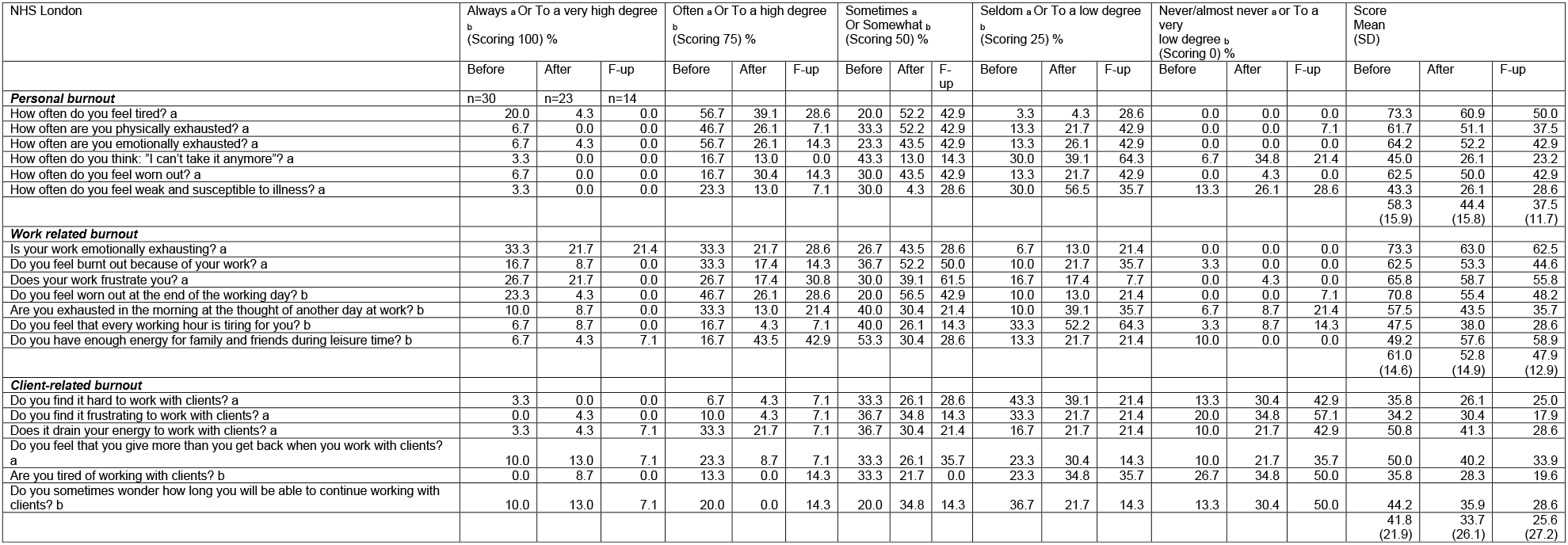
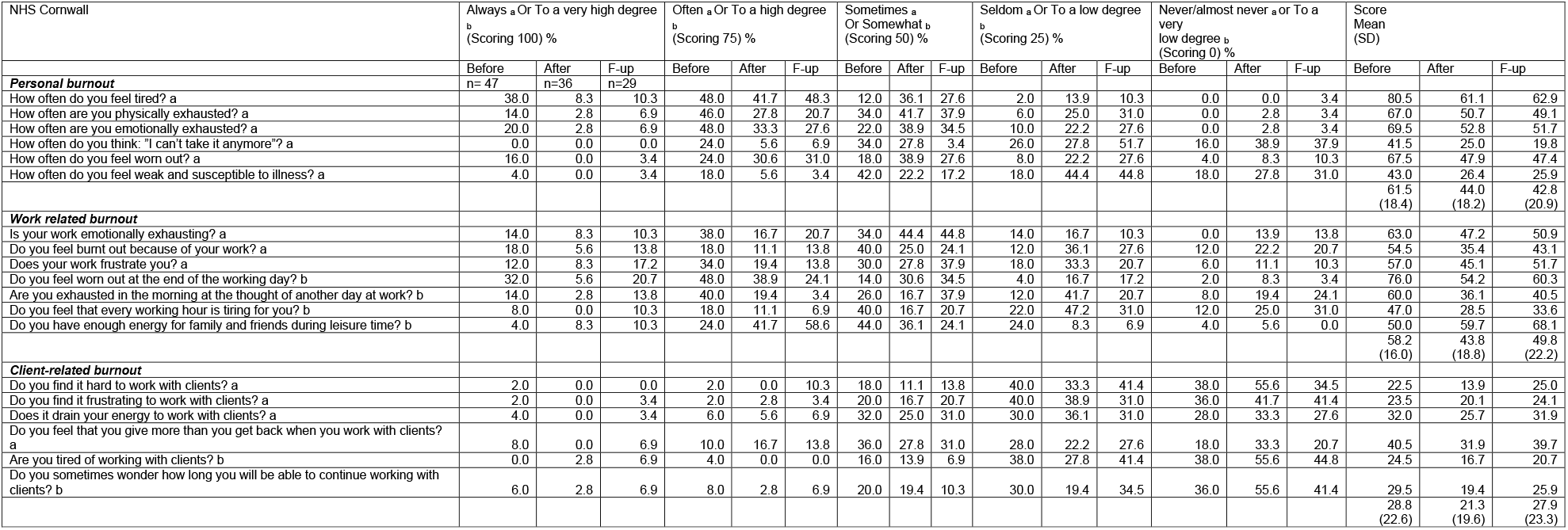

